# Image-Based Dietary Assessment Using Multimodal Large Language Models: Performance, Limitations, and Clinical Applicability in Diabetes

**DOI:** 10.64898/2026.10.21.26365949

**Authors:** Lila Gravellier, Chuyi Zhang, Yue Wu, Vijaya Parameswaran, Agatha Scheideman, Mandy Shao, Emma Zhang, Dalia Perelman, Christopher Gardner, David C. Klonoff, Helge Ræder, Michael P. Snyder, Tao Wang

## Abstract

**Background:** Image-based dietary assessment is valuable for precision diabetes care. Multimodal large language models (mLLMs) may substantially facilitate dietary self-monitoring. However, comprehensive evaluation of their real-world accuracy remains limited.

**Methods:** We developed SeeFood, an open-source benchmark, and evaluated eight commercial and open-source mLLMs on 80 real-world meal photographs from the SNAPMe dataset for nutrient and energy estimation (calories, carbohydrates, protein, fat, fiber), food-item identification, and day-level diet quality (Healthy Eating Index, HEI-2020). Three finalists were evaluated on the complete dataset (1680 photographs, 275 participant-days), and the best model was analyzed by meal type, portion size, sizing-marker use, ingredient visibility and meal complexity.

**Results:** GPT-4o mini achieved the lowest errors, with a significant advantage for carbohydrates; the three finalists performed similarly on other outcomes and on HEI-2020. Mean absolute percentage errors (MAPE) remained high (52%–90%), and the direction of bias was model-specific. The carbohydrate mean absolute error was 13.3 g (95% CI 12.4–14.3), and only 57% of meals were within ±10 g, a clinically relevant threshold for insulin dosing in type 1 diabetes. Calorie errors for lunch and dinner were in the range reported for human estimation on other images (∼55% MAPE), whereas drink-only images had the highest relative errors (312% MAPE) because of dissolved sugars. Food items were reliably identified when fully visible (Dice = 0.74), and HEI-2020 estimates showed moderate agreement with reference values (RMSE = 12.5 points; r = 0.61). Larger portions were increasingly underestimated, and sizing references did not improve accuracy. Newer or larger models and chain-of-thought prompting did not consistently improve performance.

**Conclusions:** Current mLLMs can extract meaningful dietary information from real-world meal photographs, but substantial errors in nutrient quantification and portion-size estimation limit their use for precision diabetes management. Integrating visual assessment with additional contextual information may enable more reliable, lower-burden dietary monitoring.

## 1. Introduction

Food logging, a form of dietary self-monitoring, is a cornerstone of diabetes management. In type 1 diabetes, accurate carbohydrate counting, derived from food logs, is directly linked to glycemic time-in-range^1,2^: in type 2 diabetes and prediabetes, sustained self-monitoring is associated with improved glycated hemoglobin and weight loss^3,4^. Yet food logging is burdensome, and adherence declines over time^5,6^, compromising its effectiveness in improving glycemic control and weight management outcomes.

Dietary assessment technology has evolved to reduce food-logging burden, progressing from written logs to search-based apps to image-based food recognition^7^. Multimodal large language models (mLLMs) represent the newest step in this progression. Unlike earlier computer vision methods, which were typically trained on task-specific labeled data for a single task such as food classification or portion estimation^8^, a general-purpose mLLM can in principle perform food identification, nutrient estimation, and structured output generation simultaneously, without task-specific training^9^. This capability, together with broad commercial API access and rapid ongoing development, has fueled considerable interest in mLLMs as a deployable dietary assessment tool^10,11^. However, evidence for the reliability of mLLMs in dietary assessment remains fragmented. Existing evaluations typically test a limited number of models on small samples of data curated by the model developers themselves, rather than independent benchmarks^9,12–16^. Some studies only focus on caloric accuracy, without addressing macronutrient estimation^17,18^, while others rely on lab-controlled rather than real-world images^13,16,18^. Their differences in models, datasets, and evaluation metrics hinder direct comparisons across studies. More importantly, no single study characterizes mLLM performance across the range of tasks essential to dietary assessment. These limitations highlight a need for a standardized benchmark that evaluates models against common datasets, tasks, and metrics.

To address this need, we developed SeeFood, an open-source framework for evaluating dietary-assessment accuracy of mLLMs against common data, tasks, and metrics. Notably, SeeFood tasks span nutrient and energy estimation, food identification, and diet quality, assessed using the Healthy Eating Index (HEI)–a validated measure linked to clinically relevant outcomes in diabetes^19–21^. Using SeeFood, we benchmarked representative commercial and open-source mLLMs, characterized their prediction errors, and identified their opportunities and limitations for diabetes management.

## 2. Methods

This study was designed as a retrospective benchmark of image-based dietary assessment using existing labeled datasets. Multiple multimodal large language models were evaluated and compared across three complementary levels of dietary information: meal-level nutrient and energy estimation, meal-level food item prediction, and day-level diet quality assessment. The benchmark proceeded in two phases: an initial screening evaluation across eight models on a subset of the data, followed by a full evaluation of the three best-performing models and an in-depth analysis of the best overall model (Figure 1).

**Figure 1:**
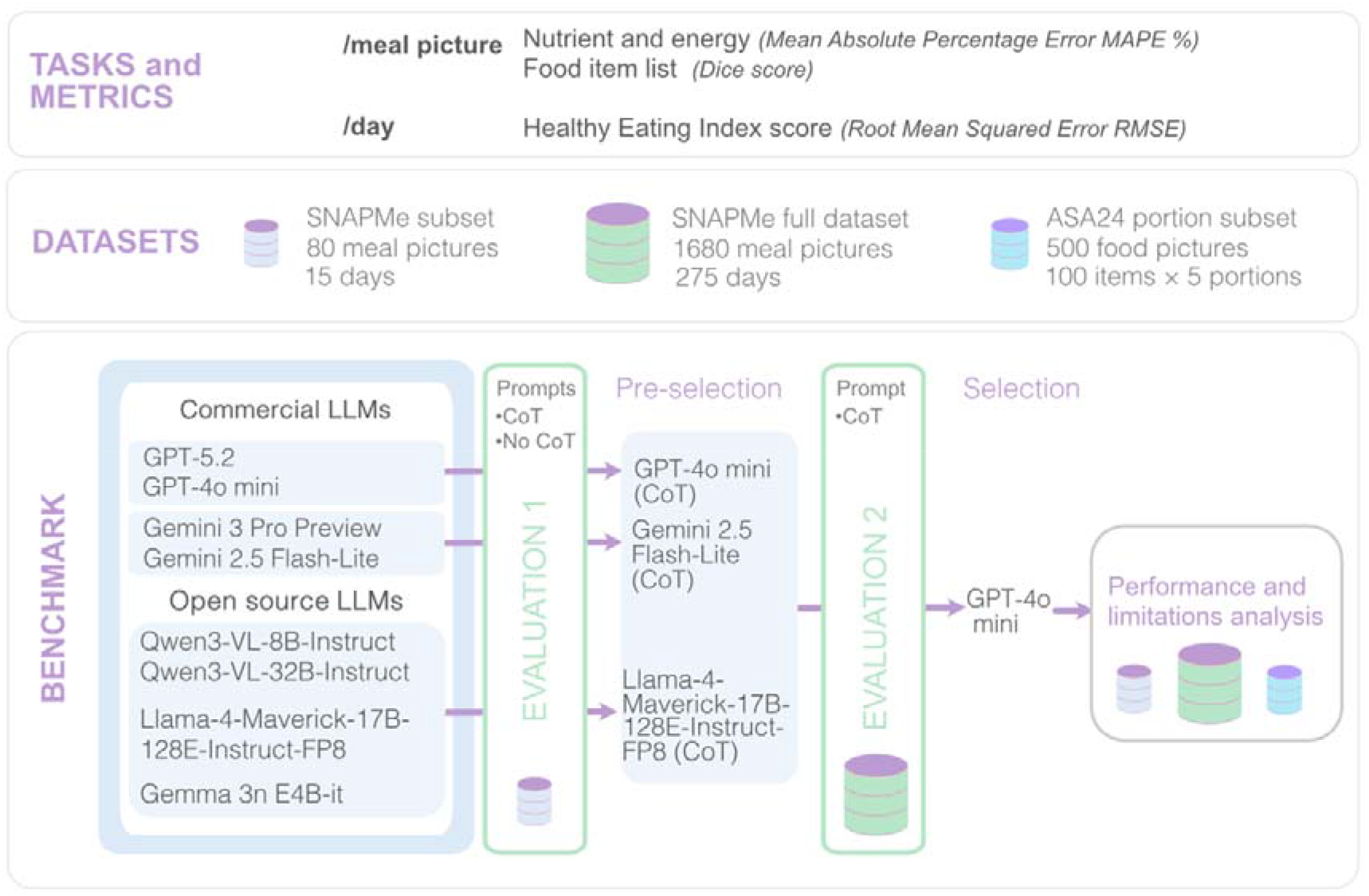
Study design and two-phase benchmark workflow for evaluating multimodal LLMs in image-based dietary assessment.

### 2.1. Evaluation tasks and metrics

Model performance was evaluated across three complementary dimensions: (1) meal-level nutrient and energy estimation, (2) meal-level food-item identification, and (3) day-level diet-quality assessment. At the meal level, the evaluated outputs were energy intake (kcal) and four nutrients: carbohydrates, protein, total fat, and fiber, as well as the list of food items present in each meal. At the day level, overall diet quality was assessed using the Healthy Eating Index-2020 (HEI-2020)^22^, a composite score ranging from 0 (poorest) to 100 (best possible diet), computed from thirteen food groups and nutrient components standardized per 1,000 kcal. The estimated HEI score was calculated from the predicted food group and nutrient portions and the predicted calories, following official NCI scoring guidelines^22^.

These three dimensions provide complementary information relevant to different clinical and dietary applications. In the context of diabetes management, meal-level nutrient estimates could inform insulin-dosing decisions; food-item identification could support actionable dietary changes, such as replacing foods high in carbohydrates with more suitable alternatives; and day-level diet-quality assessment could enable long-term dietary monitoring, help track improvements, and identify dietary patterns associated with an increased risk of complications. Evaluating each dimension separately is therefore important, because strong performance in one task does not necessarily translate into strong performance in another.

Each dimension was evaluated using metrics suited to its specific type of output. A summary of the tasks and metrics is given in Table 1, full definitions, equations and matching rules are provided in Supplementary Methods. For meal-level energy and nutrient estimates, the primary metric was the mean absolute percentage error (MAPE), which expresses error relative to the reference value independently of unit and was used to rank models. A near-zero rule was applied to reference values below 1 g or 1 kcal. Mean absolute error, root mean squared error (RMSE), mean and median signed error (bias), the proportion of meals within ±10 g of carbohydrates, and Pearson correlation were reported as secondary measures. Food-item identification was assessed with the meal-level Sørensen–Dice coefficient after name normalization and lenient one-to-one matching (e.g., predicted "rice" matched reference "brown rice"). Day-level HEI-2020 total scores (0–100 points) were evaluated primarily with RMSE. Uncertainty was quantified with participant-level bootstrap 95% CIs (2,000 resamples), retaining all photographs or participant-days of each sampled participant. Finalist models were compared using bootstrap CIs of paired differences, Bonferroni-adjusted for the two comparisons per metric (97.5% CIs). Prompt conditions in Phase 1 and the secondary analyses were compared with paired Wilcoxon signed-rank tests (p < 0.05).

**Table 1:** Evaluation tasks, definitions, clinical rationale, and performance metrics.

| Task ID | Task Name | Task Definition | Clinical Rationale | Metrics |
| --- | --- | --- | --- | --- |
| 1 | Meal-level nutrient and energy estimation | From a single meal photograph, predict energy (kcal) and four nutrients, including carbohydrate, protein, total fat, and fiber (g). | Energy and macronutrient estimation characterizes overall meal composition, with carbohydrate estimation being particularly relevant to glycemic management in diabetes<sup>1,2</sup> | Mean Absolute Percentage Error (MAPE, primary); Mean Absolute Error (MAE) in original units, Pearson r, and mean signed difference (bias). Model comparisons: participant-level bootstrap of paired differences. |
| 2 | Meal-level food item identification | From a single meal photograph, predict the list of food items present in the meal. | Food-item identification helps explain the sources of nutrient estimates<sup>23</sup> and, because dietary guidelines are expressed in terms of foods and food groups, provides actionable information to guide dietary changes<sup>24</sup> | Sørensen–Dice coefficient (primary); Model comparisons: participant-level bootstrap of paired differences. |
| 3 | Day-level diet quality assessment | From a full day of meal and drinks photographs (including breakfast, lunch, dinner, snacks, drinks), aggregate per-photograph predictions of energy and food portions to estimate the HEI-2020 total score (0-100). | The HEI-2020 captures overall dietary quality beyond individual meals and is associated with clinically relevant outcomes in diabetes<sup>19–22</sup> | Root Mean Squared Error (RMSE, primary); Pearson r. Model comparisons: participant-level bootstrap of paired differences. |

### 2.2. Datasets

#### 2.2.1. SNAPMe dataset

The Surveying Nutrient Assessment with Photographs of Meals (SNAPMe) dataset^25^ is a collection of three-day food recall from 95 US citizens. This is a publicly available benchmark dataset designed to evaluate and improve AI and computer vision models for photo-based dietary assessment^25^. Participants photographed their meals and completed Automated Self-Administered 24-Hour Dietary Assessment (ASA24) recalls, providing detailed reference nutritional information. The full dataset includes 1680 pre-meal photographs across 275 separate days. The images capture diverse real-world dietary conditions with variations in illumination, plating styles, and background objects. They include plated meals, drinks, packaged foods, and snacks. Each photograph contains a standardized 1.5×1.5-inch black-and-white paper sizing marker placed adjacent to the food. Examples of pictures from the dataset are provided in Figure 2. The SNAPMe dataset provides ground-truth labels for all three evaluation outputs: meal-level nutrient quantities, food item lists, and the dietary information required to compute day-level HEI scores.

**Figure 2:**
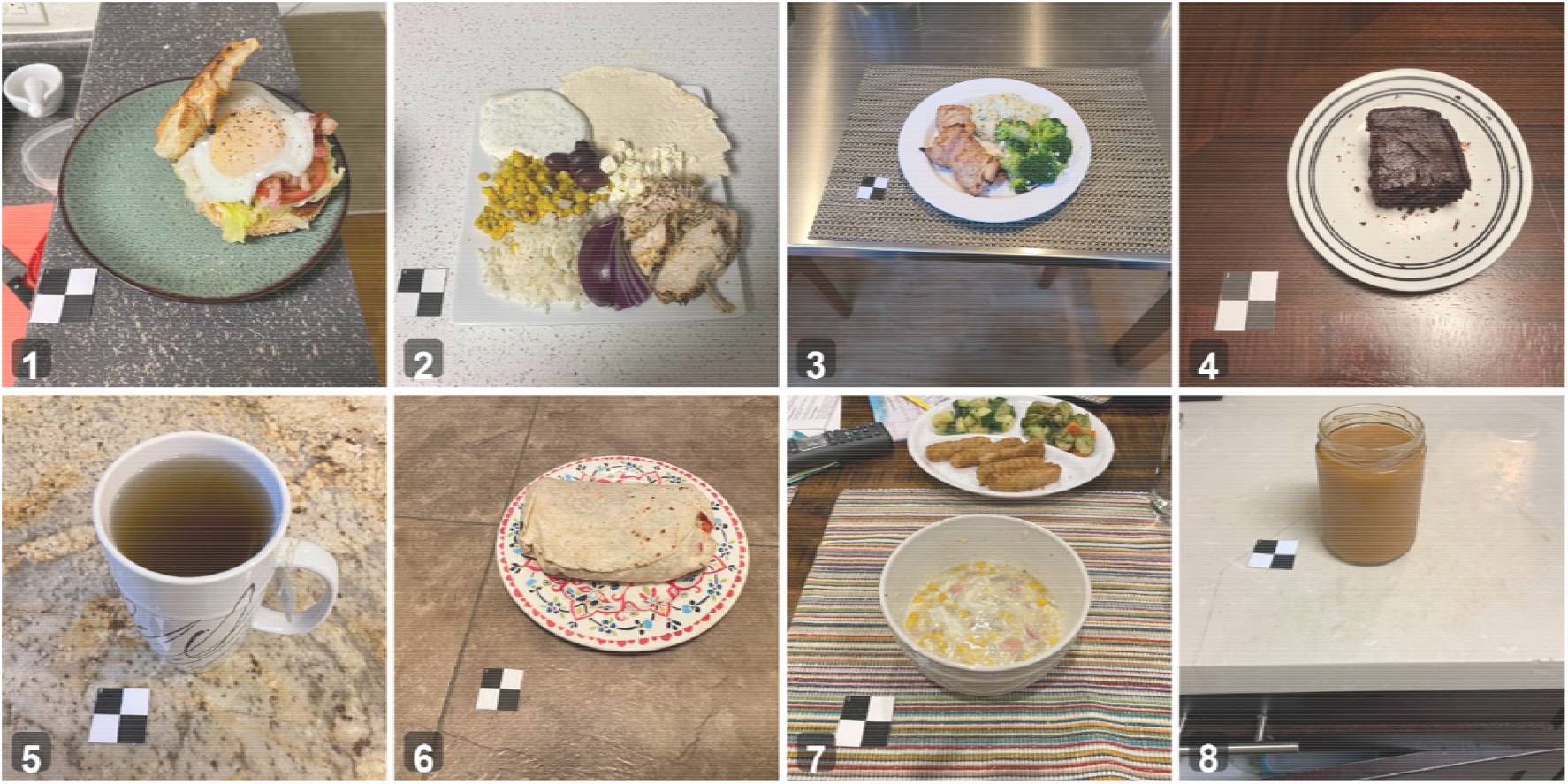
Examples of meal photographs from the SNAPMe dataset. Ground truth labels and predictions from GPT-4o mini model for these pictures are given in Table 3.

For initial model screening, we extracted a subset of SNAPMe: 15 participants were randomly selected without replacement, and their first-day food logs were retained, yielding 80 meal photographs. This subset allowed us to evaluate a larger number of models before selecting candidates for comprehensive evaluation.

#### 2.2.2. ASA24 portion dataset

To evaluate whether model outputs scale appropriately with portion size, we curated a secondary dataset by reusing a subset of ASA24 portion library^26^. This standardized image library depicts food items at multiple portion sizes, developed to visually estimate serving sizes when using the ASA24. From the full food and beverages library (n=1762), food items with images available at five portion sizes, ranging from a reference serving (1×) to five times that amount (5×) were identified (n=167). A random sample of 100 food items was selected from this subset. For each item, all five corresponding images (1×, 2×, 3×, 4×, and 5× the reference serving) were included in the analysis dataset. This subset was used exclusively for the portion sensitivity analysis described below.

### 2.3. Models and prompts

#### 2.3.1. Model selection

Eight mLLMs capable of accepting image inputs and producing structured JSON output were evaluated. Models were required to be accessible via API through OpenAI, Google AI, or TogetherAI. Commercial models included GPT-4o mini, GPT-5.2, Gemini 3 Pro Preview (hereafter Gemini 3 Pro) and Gemini 2.5 Flash-Lite. Open-source models included Qwen3-VL-8B-Instruct (Qwen3-VL 8B), Qwen3-VL-32B-Instruct (Qwen3-VL 32B), Llama-4-Maverick-17B-128E-Instruct-FP8 (Llama 4 Maverick) and Gemma-3n-E4B-it (Gemma 3n). The selection deliberately included both recent high-performance frontier models and lightweight, cost-efficient alternatives to assess the trade-off between capability and cost. Details about the different models are given in Supplementary Table S1.

#### 2.3.2. Prompt strategies

All models were required to return a structured JSON output containing ingredient- and meal-level nutrient estimates and Healthy Eating Index (HEI)–relevant portion variables, with numeric fields set to zero when not applicable. Two prompt strategies were systematically compared across all models. The first directly requested the completed JSON without explicit reasoning guidance (no-CoT). The second used structured chain-of-thought (CoT) guidance to prompt sequential ingredient identification, portion-size and nutrient estimation, meal-level aggregation, estimation of HEI-relevant variables, and a final consistency check before output generation. The same prompt text was used for all models within each condition. Additional prompt variants were evaluated in the sizing-marker sensitivity analysis. All prompts are available in Supplementary Methods SM6.

#### 2.3.3. Model execution

Images were evaluated in batches using SeeFood benchmark harness via API on the respective hosting platforms (OpenAI, Google AI, TogetherAI). The prompt was supplied in the system role and the image as user content. A default temperature of 0.0 for high reproducibility was applied when configurable. This means that the model was configured to be as deterministic as possible, and for the same image and the same input, it was intended to produce the same output rather than introducing randomness into its prediction. Each image-model combination was run three times with the same parameters. For quantitative nutrient outputs, the median across the three runs was taken as the final prediction. For food item lists, a majority-vote rule was applied: items appearing in at least two of the three runs were retained in the final output.

#### 2.3.4. Evaluation process

As illustrated in Figure 1, evaluation proceeded in two sequential phases. In the first phase (Evaluation 1), all eight models were assessed with both prompt strategies on the SNAPMe subset. Based on these results, the best-performing model-prompt combination was selected from each provider category (OpenAI, Google AI, open-source). This process yielded three finalists. In the second phase (Evaluation 2), these three models were evaluated on the full SNAPMe dataset. The best-performing model was then selected for subsequent in-depth analysis. We developed and made available this entire evaluation pipeline as an open-source package (see Section 6) to support reproducibility and facilitate its reuse to evaluate new or future models and promote strategies for additional dietary assessments.

### 2.4. Model performance analysis

#### 2.4.1. Performance by meal type

Nutrient prediction errors were stratified by meal category (breakfast, lunch, dinner, snack, drink-only) and compared, as an indicative reference, with human calorie estimation reported on a different image dataset^27^.

#### 2.4.2. Portion size sensitivity

Using the curated ASA24 subset (Section 2.2.2), model predictions for each food item were compared across increasing portion sizes. This analysis assessed whether nutrient estimates scaled with the true portion size, and in particular whether larger portions were systematically underestimated.

#### 2.4.3. Sizing marker effect

A standardized background reference object may help models in assessing portion sizes. As aforementioned, SNAPMe dataset has a 1.5×1.5-inch black-and-white paper sizing marker on each image. On our SNAPMe subset, four prompt conditions were compared: 1) standard CoT (no marker reference), 2) CoT with the true marker size disclosed, 3) CoT with an artificially larger size, and 4) CoT with an artificially smaller size. MAPE was compared across conditions.

#### 2.4.4. Food item analysis: visibility, complexity and relationship with nutrient estimation

A subset of the SNAPMe dataset was curated to understand the impact of food item visibility and complexity over model predictions. Meals were classified as fully visible (all ground-truth items visually accessible) or partially hidden (at least one item concealed). Dice scores were compared between these groups on a balanced subsample matched by the number of food items. Dice performance was also analyzed according to the ground-truth food item count, which served as a proxy for meal complexity. Finally, the correlation between the Dice and each of the per-meal nutrients and energy APE scores was calculated on the subset of images with exactly 3, 4 and 8 ground-truth food items.

#### 2.4.5 HEI component-level error analysis

To examine how portion estimation errors translated into HEI-2020 score errors, we compared predicted and reference daily food-group amounts and their corresponding component scores across the models. We inspected cases where different portion estimates yielded similar component scores, or where larger portion errors resulted in smaller component score errors. This descriptive analysis explored how energy normalization and component scoring thresholds influenced the relationship between portion accuracy and HEI score accuracy.

## 3. Results

### 3.1. Phase 1: Initial benchmark

#### 3.1.1. Model and prompt performance

Complete per-model and per-nutrient MAPE results for all 16 model-prompt combinations are reported in Supplementary Table S2. Three overarching observations emerged from this initial screening.

First, errors were substantial across all models and conditions. No model achieved a MAPE below 43% for any individual nutrient (Supplementary Table S2). For screening, we used an equally weighted composite (mean MAPE across the five outcomes), which ranged from 57.6% to 99.3%; nutrient-specific MAPEs were used for interpretation.

Second, the benefit of chain-of-thought prompting was model- and outcome-dependent and was non-significant in most comparisons. Statistical significance was assessed within each model using one-sided paired Wilcoxon signed-rank tests, testing the directional hypothesis that CoT performed better than No-CoT (p<0.05). No significant CoT benefit was observed for calories, protein, total fat, or HEI-2020 estimation in any model. Significant effects were limited to a small number of model-nutrient combinations (details in Supplementary Table S2). Overall, the benefits of CoT were observed primarily for food-item identification, with significant improvements in Dice scores for four models. Llama 4 Maverick showed the broadest benefit, with significant improvements in food-item identification, carbohydrate and fiber estimation, and mean MAPE across the five meal-level quantitative outcomes.

Third, neither model size nor recency predicted performance consistently across output types. GPT-5.2, the most recent OpenAI model at the time we ran the experiment, was outperformed by GPT-4o mini on all three evaluation dimensions (nutrient MAPE, Dice, and HEI RMSE). Within the Google family, Gemini 3 Pro achieved the lowest mean nutrient MAPE of any model in the subset (57.6%), yet Gemini 2.5 Flash-Lite outperformed it on Dice and HEI, illustrating that rankings are not stable across output types.

#### 3.1.2. Model selection

For each provider, the model-prompt combination winning the most evaluation categories among composite nutrient MAPE, Dice score, and HEI RMSE was selected for full-dataset evaluation. GPT-4o mini with CoT was selected for OpenAI (best on 2 of 3 categories), Gemini 2.5 Flash-Lite with CoT for Google (best on Dice and HEI despite higher nutrient MAPE), and Llama 4 Maverick with CoT for open-source (best on nutrient MAPE and Dice). These three finalists were carried forward to Phase 2.

### 3.2. Phase 2: evaluation of the three pre-selected models on the full SNAPMe dataset

Main results are reported in Table 2, full metrics (RMSE, correlations, median bias) in Supplementary Table S3, and error profiles in Figure 3A–C.

**Figure 3:**
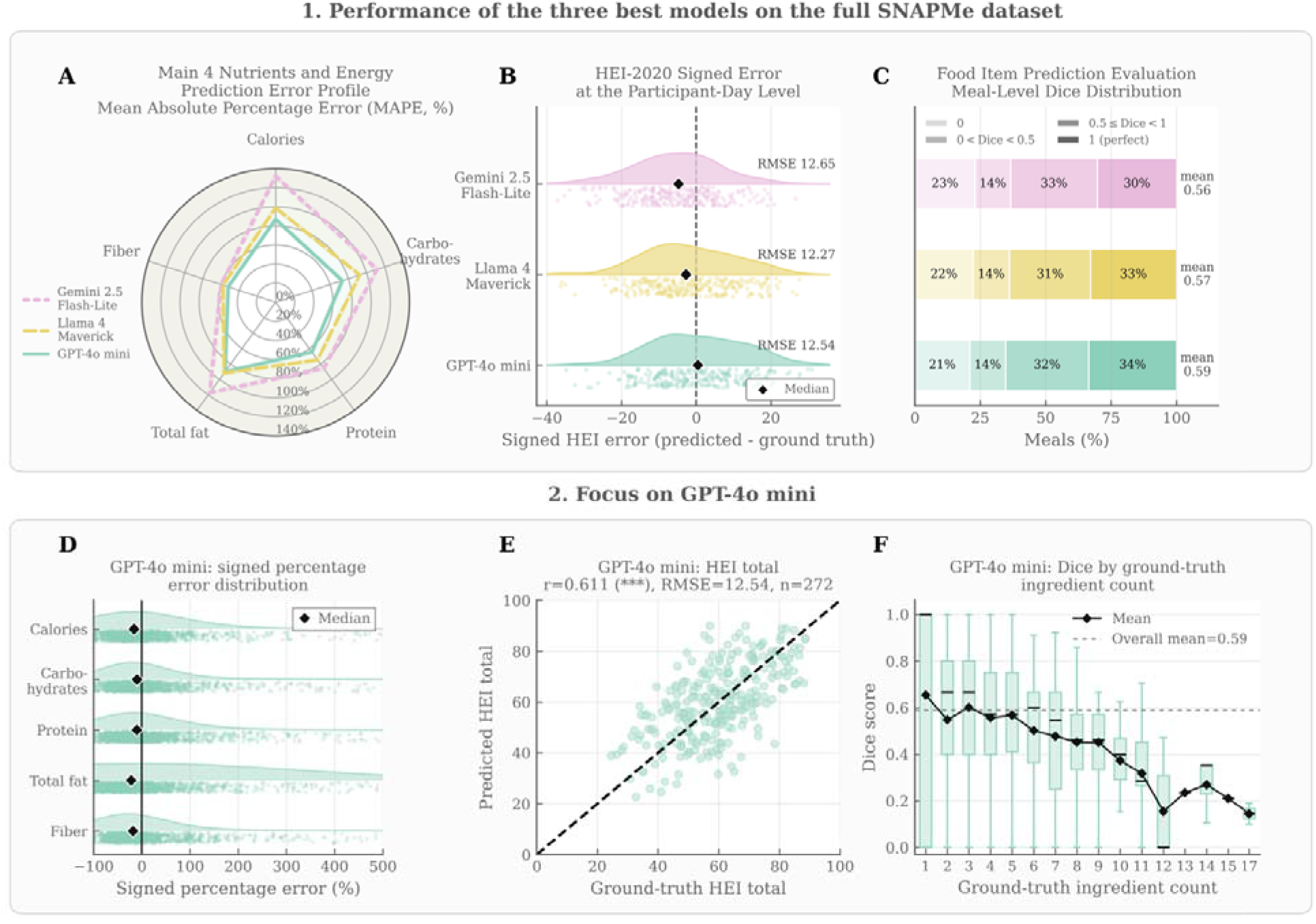
Performance of the three finalist models on the full SNAPMe dataset and error profile of GPT-4o mini. (A) MAPE by outcome. (B) Distribution of signed HEI-2020 error (predicted − reference) per participant-day (n = 272 with valid scores); diamonds, medians; RMSE shown for each model. (C) Distribution of meal-level Dice scores in four classes (0, 0 < Dice < 0.5, 0.5 ≤ Dice < 1, 1); mean Dice shown for each model. (D) Signed percentage errors of GPT-4o mini by outcome (x-axis truncated at 500%); diamonds, medians. (E) Predicted versus reference HEI-2020 total scores for GPT-4o mini; dashed line, perfect agreement; Pearson correlation, ***p < 0.001. (F) Dice score of GPT-4o mini by number of ground-truth food items; boxes, interquartile range; diamonds, means; dashed line, overall mean. Confidence intervals and model comparisons are reported in Table 2.

**Table 2:** Performance metrics of the three finalist models across nutrients, energy, HEI, and food item prediction outputs. Brackets give percentile 95% CIs from 2,000 participant-level bootstrap resamples; all photographs or participant-days from each sampled participant were retained. Positive bias indicates overestimation. Bold indicates that the best point estimate was significantly better than both other models, based on participant-level bootstrap CIs of paired differences, Bonferroni-adjusted for the two comparisons per row (97.5% CIs). RMSE, Pearson correlations, median bias and additional HEI metrics are reported in Supplementary Table S3.

| Outcome | Metric | GPT-4o<br>mini | Gemini 2.5<br>Flash-Lite | Llama 4<br>Maverick |
| --- | --- | --- | --- | --- |
| **Calories**<br>(kcal) | MAPE (%) | 86.8 [65.1, 110.9] | 132.0 [99.3, 174.4] | 98.8 [75.9, 124.3] |
|  | MAE | 123 [115, 133] | 128 [120, 137] | 130 [120, 140] |
|  | Mean bias | -61 [-74, -50] | 30 [17, 43] | -46 [-59, -34] |
| **Carbohydrates**<br>(g) | MAPE (%) | **73.4 [61.8, 87.1]** | 111.6 [87.6, 139.5] | 92.5 [74.6, 113.6] |
|  | MAE | **13.3 [12.4, 14.3]** | 15.8 [14.6, 17.0] | 14.8 [13.6, 16.0] |
|  | Mean bias | -5.0 [-6.4, -3.7] | 4.7 [3.2, 6.3] | -2.1 [-3.6, -0.7] |
| | Within $\pm 10$ g (%) | 56.8 | 53.5 | 54.5 |
| **Protein**<br>(g) | MAPE (%) | 64.6 [55.7, 75.1] | 85.6 [72.8, 100.3] | 74.6 [64.7, 85.3] |
|  | MAE | 6.1 [5.6, 6.7] | 6.4 [5.8, 7.0] | 6.2 [5.6, 6.7] |
|  | Mean bias | -2.2 [-2.8, -1.6] | 1.3 [0.7, 1.9] | -2.2 [-2.9, -1.5] |
| **Total fat**<br>(g) | MAPE (%) | 89.8 [70.8, 113.4] | 117.2 [95.3, 143.0] | 91.7 [80.5, 103.6] |
|  | MAE | 7.2 [6.7, 7.8] | 7.2 [6.7, 7.8] | 7.7 [7.1, 8.4] |
|  | Mean bias | -3.4 [-4.1, -2.6] | **0.5 [-0.3, 1.2]** | -3.1 [-3.9, -2.4] |
| **Fiber**<br>(g) | MAPE (%) | **52.1 [47.1, 58.2]** | 59.9 [53.6, 66.9] | 57.9 [52.7, 63.3] |
|  | MAE | 1.69 [1.55, 1.84] | 1.69 [1.55, 1.84] | 1.76 [1.62, 1.90] |
|  | Mean bias | -0.70 [-0.89, -0.53] | **0.15 [-0.02, 0.31]** | -0.52 [-0.71, -0.35] |
| **Food items** | Mean Dice | 0.59 [0.56, 0.61] | 0.56 [0.54, 0.59] | 0.57 [0.55, 0.60] |
| **HEI-2020**<br>(points) | RMSE | 12.5 [11.6, 13.5] | 12.7 [11.6, 13.8] | 12.3 [11.3, 13.2] |

GPT-4o mini had the lowest MAPE for all outcomes (Figure 3A) and the highest Dice score (Table 2), but its advantage was statistically supported only for carbohydrates (MAPE 73.4% vs. 111.6% and 92.5%; MAE 13.3 g vs. 15.8 g and 14.8 g) and fiber MAPE. Differences in calories, protein, fat and Dice were not significant after correction, and absolute errors for protein, fat and fiber were nearly identical across models.

HEI total score prediction was similar across the three models (RMSE 12.54, 12.27 and 12.65 points), with largely overlapping day-level error distributions centered near zero (Figure 3B) and moderate correlations with reference scores (r = 0.61–0.65; Figure 3E, Supplementary Table S3). Food-item identification was also similar across models: about one third of meals were identified perfectly (Dice = 1; 30%–34%) and about one fifth not at all (Dice = 0; 21%–23%; Figure 3C). Across outcomes, total fat showed the weakest correlations for all models (r = 0.49–0.58), whereas protein showed the strongest (r = 0.73– 0.76; Supplementary Table S3).

The direction of bias was model-specific. GPT-4o mini and Llama 4 Maverick underestimated all outcomes on average (GPT-4o mini: −61 kcal [95% CI −74 to −50] for calories and −5.0 g [−6.4 to −3.7] for carbohydrates), whereas Gemini 2.5 Flash-Lite overestimated calories (+30 kcal [17 to 43]), carbohydrates (+4.7 g [3.2 to 6.3]) and protein (+1.3 g [0.7 to 1.9]). Median signed errors were close to zero for all models, with IQRs including zero (Supplementary Table S3), indicating that mean bias was driven by a minority of large errors rather than by a shift affecting most meals. In percentage terms, error distributions were right-skewed (Figure 3D), with occasional large overestimates of items with low reference values.

GPT-4o mini was selected for the in-depth analyses because it had the lowest errors overall and a significant advantage for carbohydrates, the most clinically relevant nutrient for diabetes management.

### 3.3. Phase 3: performance and limitations of the best-performing model

Predictions of the best-performing model for the eight example photographs of Figure 2, illustrating typical successes and failure modes are shown in Table 3.

**Table 3:**
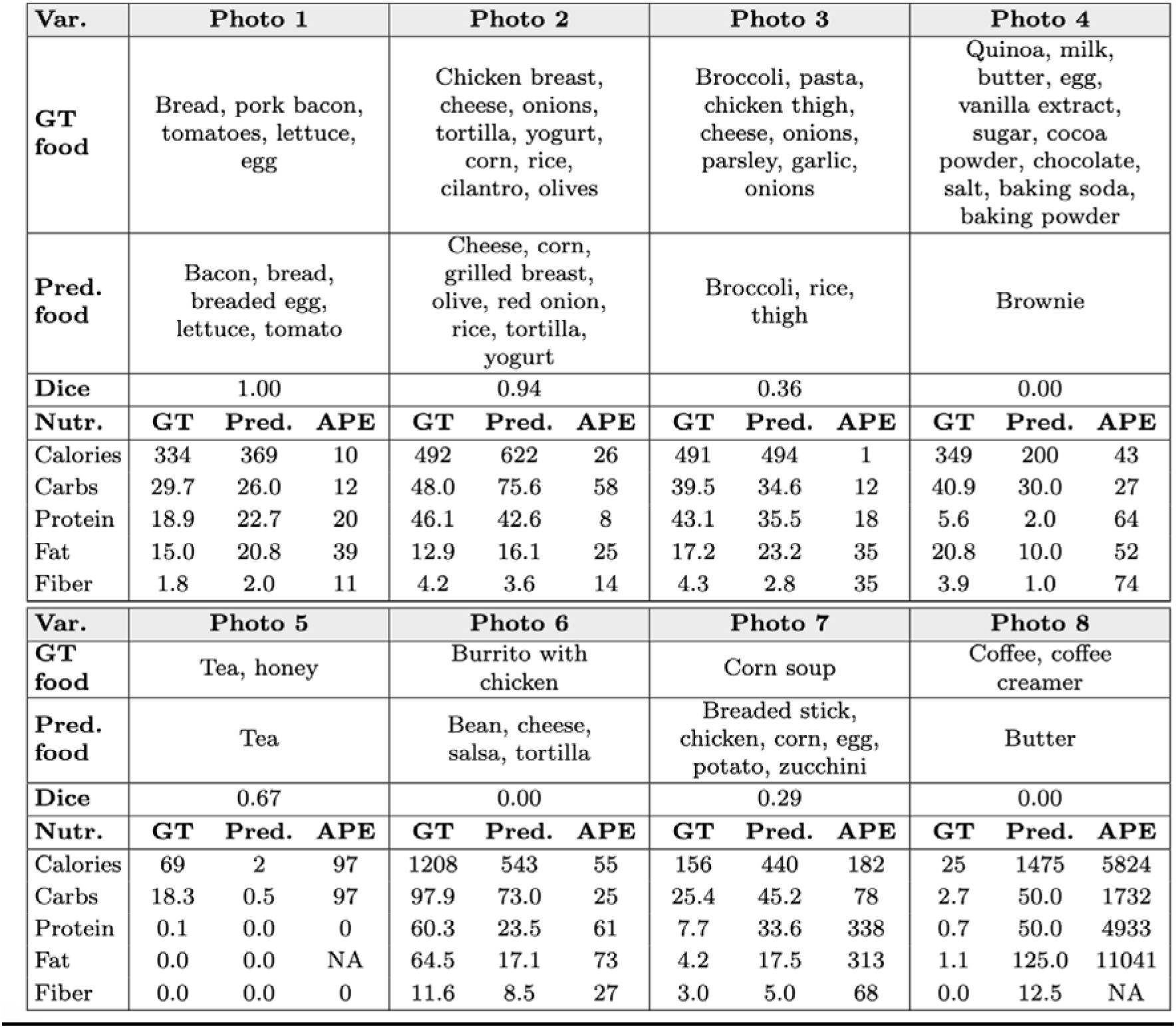
Example predictions of GPT-4o mini for eight meal photographs from Figure 2. GT, ground truth; Pred., predicted; APE, absolute percentage error (%); Dice, Sørensen–Dice coefficient for food-item identification (Supplementary Methods SM1–SM2). Energy in kcal; macronutrients and fiber in g. NA, APE undefined because the reference value is zero.

| Var. | Photo 1 |  |  | Photo 2 |  |  | Photo 3 |  |  | Photo 4 |  |  |
| --- | --- | --- | --- | --- | --- | --- | --- | --- | --- | --- | --- | --- |
| **GT food** | Bread, pork bacon, tomatoes, lettuce, egg |  |  | Chicken breast, cheese, onions, tortilla, yogurt, corn, rice, cilantro, olives |  |  | Broccoli, pasta, chicken thigh, cheese, onions, parsley, garlic, onions |  |  | Quinoa, milk, butter, egg, vanilla extract, sugar, cocoa powder, chocolate, salt, baking soda, baking powder |  |  |
| **Pred. food** | Bacon, bread, breaded egg, lettuce, tomato |  |  | Cheese, corn, grilled breast, olive, red onion, rice, tortilla, yogurt |  |  | Broccoli, rice, thigh |  |  | Brownie |  |  |
| **Dice** | 1.00 |  |  | 0.94 |  |  | 0.36 |  |  | 0.00 |  |  |
| **Nutr.** | **GT** | **Pred.** | **APE** | **GT** | **Pred.** | **APE** | **GT** | **Pred.** | **APE** | **GT** | **Pred.** | **APE** |
| Calories | 334 | 369 | 10 | 492 | 622 | 26 | 491 | 494 | 1 | 349 | 200 | 43 |
| Carbs | 29.7 | 26.0 | 12 | 48.0 | 75.6 | 58 | 39.5 | 34.6 | 12 | 40.9 | 30.0 | 27 |
| Protein | 18.9 | 22.7 | 20 | 46.1 | 42.6 | 8 | 43.1 | 35.5 | 18 | 5.6 | 2.0 | 64 |
| Fat | 15.0 | 20.8 | 39 | 12.9 | 16.1 | 25 | 17.2 | 23.2 | 35 | 20.8 | 10.0 | 52 |
| Fiber | 1.8 | 2.0 | 11 | 4.2 | 3.6 | 14 | 4.3 | 2.8 | 35 | 3.9 | 1.0 | 74 |

**Table 3: Example predictions of GPT-4o mini for eight meal photographs from Figure 2. GT, ground truth; Pred., predicted; APE, absolute percentage error (%); Dice, Sørensen–Dice coefficient for food-item**
| Var. | Photo 5 |  |  | Photo 6 |  |  | Photo 7 |  |  | Photo 8 |  |  |
| --- | --- | --- | --- | --- | --- | --- | --- | --- | --- | --- | --- | --- |
| **GT food** | Tea, honey |  |  | Burrito with chicken |  |  | Corn soup |  |  | Coffee, coffee creamer |  |  |
| **Pred. food** | Tea |  |  | Bean, cheese, salsa, tortilla |  |  | Breaded stick, chicken, corn, egg, potato, zucchini |  |  | Butter |  |  |
| **Dice** | 0.67 |  |  | 0.00 |  |  | 0.29 |  |  | 0.00 |  |  |
| **Nutr.** | **GT** | **Pred.** | **APE** | **GT** | **Pred.** | **APE** | **GT** | **Pred.** | **APE** | **GT** | **Pred.** | **APE** |
| Calories | 69 | 2 | 97 | 1208 | 543 | 55 | 156 | 440 | 182 | 25 | 1475 | 5824 |
| Carbs | 18.3 | 0.5 | 97 | 97.9 | 73.0 | 25 | 25.4 | 45.2 | 78 | 2.7 | 50.0 | 1732 |
| Protein | 0.1 | 0.0 | 0 | 60.3 | 23.5 | 61 | 7.7 | 33.6 | 338 | 0.7 | 50.0 | 4933 |
| Fat | 0.0 | 0.0 | NA | 64.5 | 17.1 | 73 | 4.2 | 17.5 | 313 | 1.1 | 125.0 | 11041 |
| Fiber | 0.0 | 0.0 | 0 | 11.6 | 8.5 | 27 | 3.0 | 5.0 | 68 | 0.0 | 12.5 | NA |

#### 3.3.1. Performance by meal type

Prediction accuracy varied substantially by meal category across all five nutrients (Figure 4A–B). Calorie MAPE was lowest for dinner (43.4%) and lunch (58.9%), and higher for breakfast (88.8%) and snacks (94.3%). Dinner nevertheless had the largest absolute error (MAE 159.3 kcal), reflecting larger portions (Supplementary Table S4). Drink-only images had the highest relative error (calorie MAPE 312.3%), indicating that the model often failed to determine whether a beverage contained energy at all, for example whether it was sweetened. Absolute errors for drinks were nevertheless small on average (MAE 48.6 kcal; 7.5 g carbohydrates; Supplementary Table S4). Two beverage examples illustrate this failure (Table 3): honey added to tea was not detected (Photo 5; 69 vs. 2 kcal), and coffee with creamer was misidentified as butter, probably because of the container (jar instead of cup), yielding 1,475 kcal instead of 25 kcal (Photo 8). As an indicative reference, nutrition experts estimating calories from a different dataset of 20 food images (12 individual food items and 8 composite dishes) showed a MAPE of ∼55%^27^; dinner and lunch errors were in this range, whereas breakfast, snacks and beverages were well above it (Figure 4A). Across nutrients (Figure 4B), lunch and dinner produced the most balanced relative errors, breakfast performed poorly for total fat (MAPE 117.4%), and beverages showed the highest relative errors for all outcomes except fiber.

**Figure 4.**
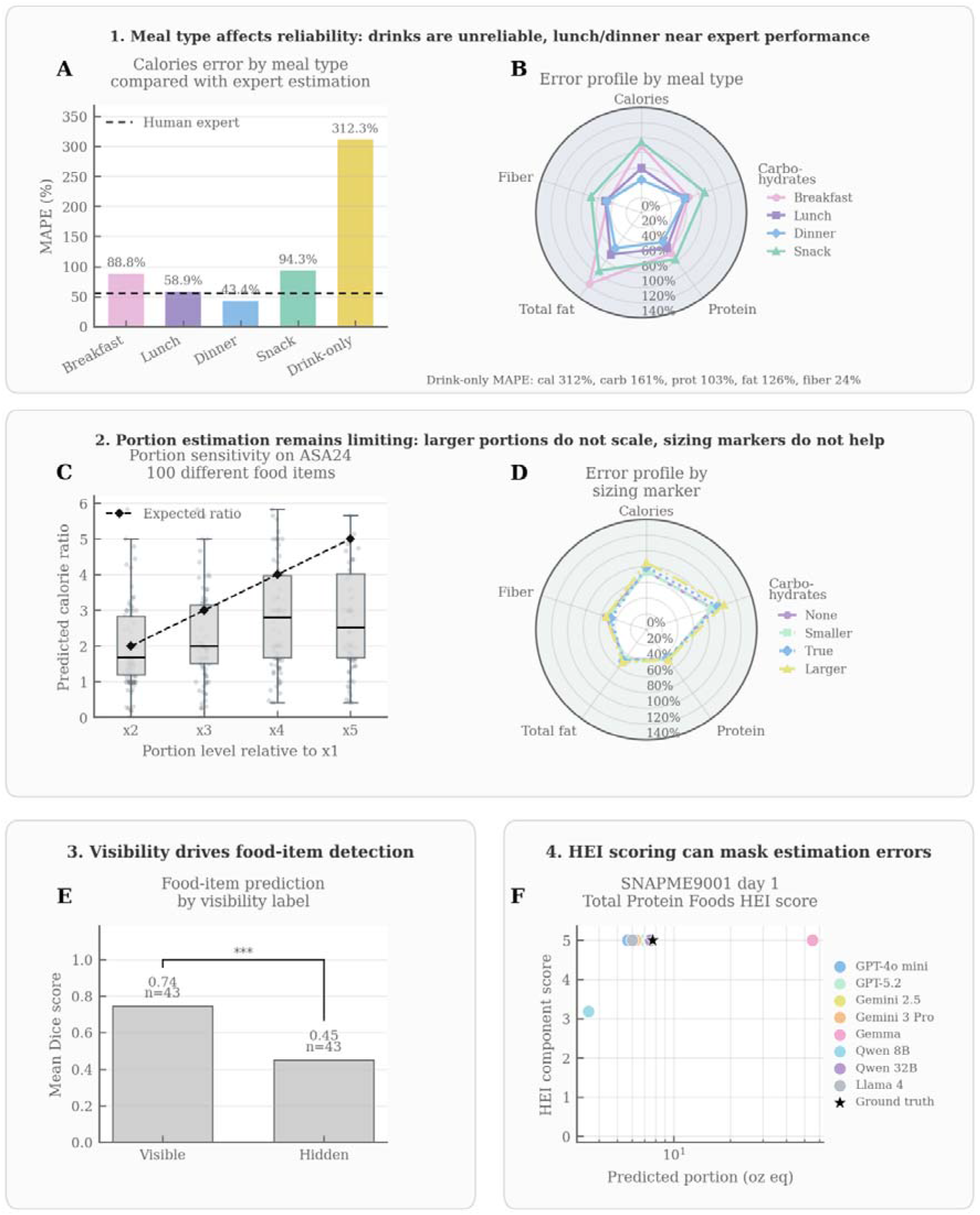
Sources of prediction error (A) Calorie MAPE by meal type; dashed line, expert human estimation on a different image set^27^. (B) MAPE by meal type across outcomes (drink-only values below the panel). (C) Predicted calorie ratio relative to the 1× portion for 100 ASA24 food items; dashed line, expected ratio. (D) MAPE by sizing-marker prompt condition (SNAPMe subset, n = 80). (E) Mean Dice score for fully visible versus partially hidden meals (n = 43 per group); ***p < 0.001. (F) Example of HEI-2020 component scoring (Total Protein Foods, one participant-day): different predicted portions can yield the same score; star, reference value (eight models, Phase 1 subset).

#### 3.3.2. Portion size sensitivity

Each image was analyzed by the model independently, without showing the model other portions of the same food for comparison. The resulting absolute predictions, when plotted against true portion size (Figure 4C), revealed a clear pattern: as the true portion increased, the model’s absolute estimates also increased, but too little, leading to underestimating larger portions. At the largest portions tested (4× and 5× the reference serving), median predicted calorie ratios plateaued at approximately 2–3×, roughly half of the true ratios (Figure 4C). This pattern held across all five nutrients (Supplementary Figure S1).

#### 3.3.3. Sizing marker effect

As shown in Figure 4D, providing explicit size information about the standardized reference marker–using its true dimensions (True) or intentionally larger (Larger) or smaller values (Smaller) produced no significant change in prediction accuracy for any nutrient relative to the baseline condition (None) in which no size information was provided in the prompt.

#### 3.3.4. Food item prediction: visibility, complexity and relationship with nutrient estimation

Item visibility was a strong determinant of food item prediction performance (Figure 4E). Mean Dice score was 0.74 when all ground-truth food items were visible versus 0.45 when at least one was hidden (n=43 per group, p<0.001). Photos 5 and 6 (Table 3) illustrate typical hidden-item meals: honey dissolved in tea and fillings concealed inside a burrito. Performance also declined steadily with meal complexity (Figure 3F), from approximately 0.6-0.7 for meals with 1-4 food items to below 0.3 for meals with 13 or more.

Despite these effects on food item identification, Dice score showed no significant correlation with nutrient estimation error for any of the five nutrients in meals with exactly 4 ground-truth food items (n=141, r=-0.035 to -0.162, all p>0.05), even when restricting to fully visible meals. This finding remained consistent when the number of food items per group was varied (see the Supplementary Figure S2 for results using three and eight food items). Table 3 illustrates this dissociation: Photo 3 had a Dice score of 0.36 but a calorie APE of 1%, whereas Photo 1 had perfect item identification (Dice = 1.00) but a fat APE of 39%. Dice also penalizes differences in naming granularity: a homemade brownie recorded as ten ingredients but predicted as "brownie" scored 0.00, although calories were within 43% (Photo 4, Table 3).

#### 3.3.5. Exceptional cases with HEI

Food component-level inspection of HEI predictions revealed cases where models with different portion estimates received identical or near-identical component scores, and cases where a model with an excessively larger absolute deviation from the true portion received a better component score than a model with a smaller deviation (Figure 4F).

## 4. Discussion

This independent benchmark evaluated eight commercial and open-source mLLMs on real-world meal photographs across three complementary outputs: energy, macronutrients and fiber, food-item identification, and day-level diet quality (HEI-2020). Current mLLMs extracted meaningful dietary information, but with errors that still prevent unsupervised clinical use. Beyond benchmarking models, the study identifies where and why errors arise, namely hidden or dissolved ingredients, ambiguous framing, meal complexity and portion size. It translates these findings into concrete guidance for prompting, image capture and evaluation metrics.

### 4.1 Main findings

For GPT-4o mini, which had the lowest errors overall and was significantly better for carbohydrates, absolute errors remained large (MAPE > 50% for all outcomes), although predictions were associated with reference values for all nutrients (r = 0.53–0.75). Food items were reliably identified when all were visible (Dice = 0.74), and the day-level HEI-2020 score was estimated with an RMSE of about 12 points (r = 0.61). This is consistent with the moderate agreement reported for other surrogate measures of diet quality^28^. Errors for lunch and dinner, however, approached published estimates for expert human calorie estimation (Zhou et al., 2018). Protein and fiber were estimated more accurately than fat, carbohydrates and energy in all three selected models, suggesting a general property of current mLLMs rather than of a single model. In contrast, the direction of bias differed between models, so bias correction cannot be generalized across models. Newer or larger models did not necessarily perform better: in the Phase 1 screening (n = 80), GPT-5.2 was outperformed by GPT-4o mini on all three evaluation dimensions and rankings within the Google family varied across outputs. Chain-of-thought prompting brought little benefit.

### 4.2 Sources of error and their implications

The largest errors arose from information that is not visible in the image. The energy content of beverages depends mainly on dissolved sugar, honey or alcohol, and concealed fillings, cooking fats and background foods create comparable ambiguity (Photo 5-6-7, Table 3). These failures are structural and might drive the right-skewed error distributions (Figure 3D) and are unlikely to be solved by generic prompt engineering. Practical mitigations are targeted follow-up questions (e.g., on added sugar, alcohol, fats or fillings) and photographing only the food being logged. To avoid a return to a burdensome manual logging, these questions should be quick to answer, appear only for high-risk items such as beverages or for added fat, and could be stored as individual habits (e.g., usual sugar in coffee) so that they are not asked repeatedly.

The decline in food item identification with meal complexity reflects more hidden ingredients but also granularity mismatches inherent to open-ended outputs. A closed food vocabulary, tailored to the individual, population or cuisine, would reduce such mismatches. Linking identified foods to a verified nutrient database, as implemented in DietAI24^16^, is complementary. It can improve the nutrient composition assigned to correctly identified foods and make estimates traceable. However, it cannot correct errors in portion size or in characteristics that are not visually observable. The absence of correlation between Dice and nutrient error might reflect a limitation of the metric itself: Dice weights all items equally, whereas a missing lettuce leaf and a missing spoonful of oil have very different nutritional consequences. Nutrient-weighted food identification metrics would better align identification with quantification.

Estimates did not change when the prompt gave the true dimensions of the reference marker, or even deliberately false ones. This suggests that models may not use the scale information provided and instead estimate quantities from the typical size of each recognized food item, and food item standard portion, which may also explain the underestimation of large portions. Improving portion estimation will likely require portion-varied fine-tuning or depth and volumetric reasoning; until then, users should photograph single standard servings rather than big servings or whole dishes, which is a major practical limitation as many real-world meals do not match a standard serving. Finally, the similar HEI RMSEs across models despite different nutrient accuracy reflect plateau effects in HEI-2020 component scoring, so HEI evaluation should include component-level accuracy.

### 4.3 Comparison with prior work and contribution

Our findings agree with recent studies. O’Hara et al.^9^ also observed increasing underestimation of larger portions, and Johansen et al.^15^ found that a size reference did not improve carbohydrate estimation for composite meals. Firdolfsson et al.^13^ reported lower errors for ChatGPT-4o on standardized laboratory images than we observed on real-world photographs (calorie MAPE 35.8% vs. 86.8%), which may suggest that standardized image acquisition (e.g., a plain background, good lighting and fully visible food) improves performance. A recent study likewise found no benefit of chain-of-thought on a SNAPMe subset^29^. Unlike their observation that newer Anthropic models performed better, newer models were not consistently better in our benchmark. Model selection for dietary assessment therefore requires task-specific evaluation rather than reliance on general-purpose benchmarks, and a cheaper model such as GPT-4o mini may match or outperform costlier ones.

Beyond confirming these patterns, this study contributes an independent, open-source benchmark spanning eight commercial and open-source models on real-world photographs, and to our knowledge is the first to jointly assess energy, three macronutrients plus fiber, food-item identification, and day-level diet quality. By localizing the mechanisms behind the errors, it offers concrete guidance for data, prompting, metric design, and system development.

### 4.4 Implications for use

Current mLLMs are not reliable enough for precision diabetes management. The carbohydrate MAE of 13.3 g (95% CI 12.4–14.3) exceeds the ±10 g threshold beyond which insulin dosing may be compromised^1,30^, and 43% of meals fell outside this range, with errors in both directions for all models: overestimation risks insulin overdosing and hypoglycemia, and underestimation risks hyperglycemia. Portion insensitivity adds further risk. The models may be useful in lower-stakes settings. In research, they could serve as a low-burden complement to dietary recalls, where meal-level errors partly average out across days^31^. They could also support qualitative food-group feedback in counseling and the tracking of dietary change over time. These uses remain to be validated, as this benchmark assessed cross-sectional accuracy and not the ability to detect within-person changes over time. The nutrients that matter most will depend on the clinical application, and performance on the five outputs evaluated here does not necessarily generalize to other clinically relevant nutrients. Saturated fat, for example, is central to cardiovascular risk management but is likely to be particularly difficult to estimate from an image, because the type and amount of cooking fat are rarely visible. Sodium and added sugars raise similar concerns.

### 4.5 Limitations

SNAPMe includes only US participants, so performance may differ for other diets, food preparations and plating conventions, and its reference values derive from ASA24 self-reported recalls, which carry their own measurement error. Model screening relied on a subset of 80 photographs (first recorded day of 15 participants), which may have influenced which models advanced to the full evaluation. The metrics also have known weaknesses: MAPE is inflated when reference values are near zero, and Dice ignores the specificity, portion size and nutritional weight of items. All models used the same prompt without model-specific optimization and photographs were evaluated without any context such as meal timing, user history or dietary restrictions. Finally, although the eight models were chosen to span commercial and open-source options of different sizes, the field is evolving rapidly, and rankings reflect the versions available at the time of evaluation.

### 4.6 Future directions

Future work should address three areas. On the modeling side, priorities include better portion-size estimation, empirically optimized follow-up questions for beverages and hidden items, and closed food vocabularies, tailored to the culture or individual and linked to nutrient databases. On the evaluation side, a human baseline, with dietitians performing the same tasks on the same images, is needed to place mLLM performance in context, together with nutrient-weighted and component-level metrics; the SeeFood benchmark could also be extended with larger and more diverse datasets and a live leaderboard evaluating models as integrated systems. Finally, validation should extend to non-US diets, multi-image contextualized inputs and prospective clinical settings, and longitudinal studies should test whether mLLM-based estimates track within-person changes in intake and diet quality, a prerequisite for dietary monitoring.

## 5. Conclusion

Current mLLM-based food image logging can extract meaningful dietary information from real-world meal photographs, but substantial errors in nutrient and portion-size estimation limit applications requiring precise quantification, particularly high-stakes applications such as carbohydrate estimation for insulin dosing in T1D. Food identification and broader measures of dietary quality may support lower-stakes dietary monitoring, although their utility for longitudinal monitoring and clinical counseling requires further validation. Improving portion estimation and incorporating contextual information about visually inaccessible ingredients will be important for future applications.

## 6. Institutional Review Board Statement

Institutional Review Board approval was not sought because this study did not involve human subjects research as defined under applicable institutional and regulatory guidelines.

## Supporting information

Supplementary results

## Data Availability

SeeFood benchmark code is open-sourced and will be made available at publication of the article. Datasets curated are also available in the form of meta data identifying the image IDs in the SNAPMe dataset. For the original image data, refer to https://agdatacommons.nal.usda.gov/articles/dataset/SNAPMe_A_Benchmark_Dataset_of_Food_Photos_with_Food_Records_for_Evaluation_of_Computer_Vision_Algorithms_in_the_Context_of_Dietary_Assessment/24856449?file=44532971

## 7. Code and Data Availability

SeeFood benchmark code is open-sourced and will be made available at publication of the article. Datasets curated are also available in the form of meta data identifying the image IDs in the SNAPMe dataset.

## 8. Author Contributions

LG: Conceptualization and study design, Data curation, Formal analysis, Investigation, Methodology, Software, Visualization, Writing – original draft, Writing – review and editing. CZ: Conceptualization and study design, Data curation, Formal analysis, Writing – review and editing. YW: Methodology, Writing – review and editing. VP: Methodology, Writing – review and editing. AS: Investigation, Writing – review and editing. MSh: Investigation, Writing – review and editing. EZ: Writing – review and editing. DP: Methodology, Writing – review and editing. CG: Conceptualization, Methodology, Writing – review and editing. DCK: Conceptualization, Supervision, Writing – review and editing. HR: Conceptualization, Supervision, Writing – review and editing. MPS: Conceptualization, Supervision, Writing – review and editing. TW: Conceptualization and study design, Methodology, Project administration, Supervision, Writing – review and editing.

## 9. Funding Acknowledgements

Dr. Yue Wu is supported by the American Diabetes Association Grant 11-23-PDF-76 (Y.W.). Dr. Vijaya Parameswaran is supported by NHLBI training grant #5T32HL16127002.

