## Supplementary results for "Image-Based Dietary Assessment Using Multimodal Large Language Models: Performance, Limitations, and Clinical Applicability in Diabetes"

**Supplementary methods**

Definitions and equations for all reported evaluation outcomes

For each quantitative outcome, $i$ indexes paired observations with finite reference and predicted values; $n$ is the number of such pairs. Meal-level outcomes are energy (kcal), carbohydrates, protein, total fat, and fiber (g). Day-level HEI-2020 scores are evaluated on participant-days with both scores available. Unless stated otherwise, a lower error denotes better performance.

**SM1. Meal-level energy and nutrient errors**

Let $G_{i}$ and $P_{i}$ be the reference and predicted quantity for photograph $i$, and let $e_{i}=P_{i}-G_{i}$. Absolute error (AE) and signed error are defined as follows:

$$\mathrm{AE}_{i}=\left| P_{i}-G_{i} \right|, e_{i}=P_{i}-G_{i}$$

Absolute percentage error (APE) uses the following near-zero rule for the five meal-level quantitative outcomes:

$$\mathrm{APE}_{i} \left( \% \right)=\left\{ \begin{matrix} 100\times\frac{\left| P_{i}-G_{i} \right|}{G_{i}}, & \text{if }G_{i}\geq1 \\ 0, & \text{if }0<G_{i}<1\text{ and }P_{i}\leq1 \\ 100\times\left| P_{i}-G_{i} \right|, & \text{if }0<G_{i}<1\text{ and }P_{i}>1 \end{matrix} \right.$$

Observations with $G_{i}=0$ have undefined APE and are omitted from MAPE, while remaining eligible for AE, MAE, RMSE, bias, and Pearson correlation. The threshold of 1 uses the outcome’s original unit (1 kcal or 1 g).

Mean absolute percentage error (MAPE) is the arithmetic mean of valid per-photograph APE values for an outcome:

$$\mathrm{MAPE} \left( \% \right)=\frac{1}{n_{+}}\sum_{i=1}^{n_{+}} \mathrm{APE}_{i} \left( \% \right), \text{over the }n_{+}\text{ observations with }G_{i}>0$$

For Phase 1 screening, a composite mean MAPE was calculated as the unweighted mean of the five separately calculated MAPEs. Because reference-value distributions differ between nutrients, this composite was used for screening only, and nutrient-specific MAPEs were used for interpretation:

$$\text{Mean MAPE}=\frac{1}{5}\left( \mathrm{MAPE}_{\mathrm{kcal}}+\mathrm{MAPE}_{\mathrm{carb}}+\mathrm{MAPE}_{\mathrm{prot}}+\mathrm{MAPE}_{\mathrm{fat}}+\mathrm{MAPE}_{\mathrm{fiber}} \right)$$

Secondary error measures retain the original unit:

$$MAE=\frac{1}{n}\sum_{i=1}^{n} \left| e_{i} \right|, RMSE=\sqrt{\frac{1}{n}\sum_{i=1}^{n} e_{i}^{2}}, bias=\frac{1}{n}\sum_{i=1}^{n} e_{i}$$

Positive bias denotes overestimation; negative bias denotes underestimation. The standard deviation of signed errors, when reported, is the sample standard deviation:

$$\mathrm{SD}\left( e \right)=\sqrt{\frac{\sum_{i=1}^{n} \left( e_{i}-bias \right)^{2}}{n-1}}$$

The median signed error is reported with its interquartile range (IQR). For carbohydrates, the proportion of meals with an absolute error of at most 10 g is also reported.

Pearson correlation describes linear association, not agreement or error magnitude:

$$r=\frac{\sum_{i=1}^{n} \left( G_{i}-\bar{G} \right)\left( P_{i}-\bar{P} \right)}{\sqrt{\sum_{i=1}^{n} \left( G_{i}-\bar{G} \right)^{2} \sum_{i=1}^{n} \left( P_{i}-\bar{P} \right)^{2}}}$$

Pearson $r$ is undefined when fewer than two paired values are available or either series has zero variance.

**SM2. Meal-level food-item identification**

Reference and predicted food-item names were normalized before matching. Text was lowercased, non-alphanumeric punctuation was removed, predefined generic descriptors were discarded, and simple rule-based singularization was applied. Ingredients were initially separated using commas, semicolons, slashes, or vertical bars. A predicted item was considered compatible with a reference item if either normalized name contained the other or if the two normalized names shared at least one token. Matching used a greedy one-to-one procedure: predicted items were examined in their original order and assigned to the first compatible, previously unmatched reference item. Consequently, each predicted and reference item could contribute to at most one true positive (TP). Let $N_{p}$ and $N_{g}$ denote the numbers of nonempty normalized predicted and reference items, respectively. False positives are $FP=N_{p}-TP$ and false negatives are $FN=N_{g}-TP$.

$$Dice=\frac{2 \mathrm{TP}}{N_{p}+N_{g}}=\frac{2 \mathrm{TP}}{2 TP+FP+FN}$$

The meal-level Dice score is 1 if both lists are empty. If exactly one list is empty, Dice is 0. Mean Dice is the arithmetic mean of meal-level Dice scores. The sample standard deviation of those scores is reported as “Std Dice” where applicable. Precision is TP / (TP + FP) and recall is TP / (TP + FN).

**SM3. Day-level HEI-2020 error**

The HEI-2020 total score (0–100 points) was computed from predicted daily food-group and nutrient amounts and predicted energy, using the same scoring framework as for reference values. The total is the sum of the 13 HEI component scores. Component scoring follows the published HEI-2020 standards; most components are energy-density based, while fatty acids use a ratio and added sugars and saturated fat use percentages of energy. The HEI total-score error is evaluated in points, not as a percentage.

$$\mathrm{HEI}_{\mathrm{total}}=\sum_{c=1}^{13} \text{component score}_{c}, h_{j}=\mathrm{HEI}_{j}^{\mathrm{pred}}-\mathrm{HEI}_{j}^{\mathrm{ref}}$$

$$\mathrm{RMSE}_{\mathrm{HEI}}=\sqrt{\frac{1}{m}\sum_{j=1}^{m} h_{j}^{2}}, \mathrm{MAE}_{\mathrm{HEI}}=\frac{1}{m}\sum_{j=1}^{m} \left| h_{j} \right|$$

Here $j$ indexes $m$ participant-days with paired scores (272 of 275 participant-days in the full dataset). Pearson $r$ for HEI is calculated from the paired daily total scores using the formula in SM1. Component-score and food-group comparisons in the error analysis are descriptive and should not be interpreted as independent validation of diet quality.

**SM4. Portion-size and subgroup analyses**

In the controlled portion series, each food has a 1× reference portion and larger levels (2×–5×). For level $k$ of food $f$ and outcome $v$, the predicted portion-response ratio is:

$$\hat{R}_{f,k,v}=\frac{P_{f,k,v}}{P_{f,1,v}}, \text{with expected ratio }R_{f,k}=k$$

Ratios require a finite, positive predicted value at 1×. For each level, the ratio error, absolute ratio error, and relative ratio error are:

$$\text{ratio error}=\hat{R}_{f,k,v}-k, \text{absolute ratio error}=\left| \hat{R}_{f,k,v}-k \right|$$

$$\text{relative ratio error} \left( \% \right)=100\times\frac{\left| \hat{R}_{f,k,v}-k \right|}{k}$$

These ratios measure scaling with portion size, not the absolute accuracy of the 1× estimate.

Subgroup MAPE and mean Dice use the same definitions as above, restricted to eligible observations in the subgroup. The association of food-item agreement with nutrient error is Pearson $r$ between per-meal Dice and per-meal APE within the specified ground-truth food-item-count stratum.

**SM5. Statistical comparisons**

CoT and No-CoT conditions were paired by photograph for meal-level outcomes and by participant-day for HEI. Paired Wilcoxon signed-rank tests (zero_method="wilcox") compared per-image APE for each nutrient, per-image mean valid APE across the five outcomes, per-image Dice, and per-day squared HEI total-score error. Undefined APE values arising from a zero reference value were excluded.

Wilcoxon tests were used for Phase 1 and secondary analyses. Directional tests assessed whether CoT produced lower nutrient and HEI errors or higher Dice than No-CoT. Statistical significance was defined as p < 0.05; these comparisons were exploratory and not corrected for multiple comparisons. Aggregate MAPE, Dice, and HEI RMSE were reported as summary measures but were not entered directly into the tests.

For the three finalist models (Phase 2), 95% confidence intervals were obtained from 2,000 participant-level bootstrap resamples (seed 42), retaining all photographs or participant-days of each sampled participant. Metrics were recalculated within each replicate. Models were compared using percentile CIs of paired differences computed within each replicate, Bonferroni-adjusted for the two comparisons against the best model per metric (97.5% CIs); a difference was considered significant when this CI excluded zero. Median bias is reported with its IQR and was not bootstrapped.

**SM6. Prompts**

[SeefoodPrompt ing_nut_hei no CoT]

You are a meticulous nutrition analyst. Given a photo of a meal, output the ingredients, nutritional values, food category estimation.

RETURN ONLY VALID JSON MATCHING THIS SCHEMA EXACTLY

{

"packaged_food_status": "fully_packaged" | "partially_packaged" | "unpackaged",

"ingredients": [

{

"name": "string",

"weight_g": number,

"nutrients": {

"calories_kcal": number,

"fat_g": number,

"carbohydrates_g": number,

"protein_g": number,

"fiber_g": number,

"added_sugars_g": number,

"sodium_mg": number

"monounsaturated_fat_g": number,

"polyunsaturated_fat_g": number,

"unsaturated_fat_g": number,

"saturated_fat_g": number,

}

}

],

"totals": {

"calories_kcal": number,

"fat_g": number,

"carbohydrates_g": number,

"protein_g": number,

"fiber_g": number,

"added_sugars_g": number,

"sodium_mg": number

"monounsaturated_fat_g": number,

"polyunsaturated_fat_g": number,

"unsaturated_fat_g": number,

"saturated_fat_g": number,

},

"hei_components": {

"total_fruit_portion_cup": number,

"whole_fruit_portion_cup": number,

"citrus_melons_berries_whole_fruit_portion_cup": number,

"other_whole_fruits_portion_cup": number,

"fruit_juice_portion_cup": number,

"total_vegetables_portion_cup": number,

"vegetable_legumes_portion_cup": number,

"total_vegetables_and_legumes_portion_cup": number,

"dark_green_vegetables_portion_cup": number,

"greens_and_beans_portion_cup": number,

"whole_grains_portion_oz": number,

"refined_grains_portion_oz": number,

"dairy_portion_cup": number,

"meat_poultry_seafood_protein_portion_oz": number,

"egg_protein_portion_oz": number,

"nuts_and_seeds_protein_portion_oz": number,

"soy_protein_portion_oz": number,

"legume_protein_portion_oz": number,

"high_fat_seafood_protein_portion_oz": number,

"low_fat_seafood_protein_portion_oz": number,

"total_protein_foods_portion_oz": number,

"seafood_and_plant_proteins_portion_oz": number,

}

}

[Seefood ing_nut_hei CoT]

You are a meticulous nutrition analyst.

INPUT: One photo of a meal (45-degree to top-down). The meal may be hot or cold, mixed plate, snack, drink, and may include packaged foods.

GOAL: Strictly (1) identify visible ingredients, (2) estimate weights, (3) estimate typical nutrient composition, (4) compute totals, and (5) estimate HEI-relevant portions.

You DO NOT compute Healthy Eating Index (HEI) scores. You only output the HEI-related portion variables specified below.

CRITICAL RULES

- Do NOT invent ingredients not supported by visible evidence.

- When uncertain, use conservative, typical serving-size estimates.

- Use canonical, generic ingredient names (e.g., "chicken breast", "white rice", "broccoli"), not brands unless clearly visible on packaging.

- Assume edible portion only (exclude bones, peels, shells) unless clearly being consumed.

- You do not have access to external databases. Nutrient values must be estimated using standard reference-like averages (USDA-style typical values).

- Return ONLY valid JSON (no markdown, no prose, no comments).

- All fields must be present. If not applicable or not present, output 0.

OUTPUT PRECISION / ROUNDING

- weight_g: 1 decimal

- calories_kcal: nearest integer

- nutrients in grams: 1 decimal

- sodium_mg: nearest integer

- ratios: 2 decimals

- All outputs must be numeric (use 0 rather than null).

WORKFLOW (follow exactly)

1) Identify all visible ingredients.

- If the meal includes packaged items, detect whether the overall meal is: "fully_packaged", "partially_packaged", or "unpackaged".

- Do not list packaging as an ingredient.

2) Estimate the weight in grams (weight_g) for each ingredient.

3) For each ingredient, estimate typical nutrients per 100 g for:

calories (kcal), fat (g), carbohydrates (g), protein (g), fiber (g), fatty_acids (g), saturated_fats (g), added_sugars (g), sodium (mg).

Then SCALE these per-100-g values to the estimated ingredient weight.

In the JSON output, report ONLY the SCALED nutrient values for each ingredient (not the per-100-g values).

4) Compute meal totals by summing the scaled nutrient values across all ingredients.

5) Estimate HEI-relevant portions for the meal. You are NOT calculating an HEI score.

- Output portions in the correct unit (cups, ounce equivalents, or grams depending on the variable).

- Use standard USDA-style assumptions when needed (e.g., 1 cup raw leafy greens = 0.5 cup equivalent).

- If an ingredient does not contribute to a category, set that category contribution to 0.

- All HEI fields must be present.

HEI COMPONENT RELATIONSHIPS

HEI_component_structure = {

"total_fruit_portion_cup":

"citrus_melons_berries_whole_fruit_portion_cup + other_whole_fruits_portion_cup + fruit_juice_portion_cup",

"whole_fruit_portion_cup":

"citrus_melons_berries_whole_fruit_portion_cup + other_whole_fruits_portion_cup",

"total_vegetables_and_legumes_portion_cup":

"total_vegetables_portion_cup + vegetable_legumes_portion_cup",

"greens_and_beans_portion_cup":

"dark_green_vegetables_portion_cup + vegetable_legumes_portion_cup",

"whole_grains_portion_oz":

"portion of whole grain ounce equivalents",

"refined_grains_portion_oz":

"portion of refined grain ounce equivalents",

"dairy_portion_cup":

"portion of dairy in cup equivalents",

"total_protein_foods_portion_oz":

"meat_poultry_seafood_protein_portion_oz + egg_protein_portion_oz + nuts_and_seeds_protein_portion_oz + soy_protein_portion_oz + legume_protein_portion_oz",

"seafood_and_plant_proteins_portion_oz":

"high_fat_seafood_protein_portion_oz + low_fat_seafood_protein_portion_oz + soy_protein_portion_oz + nuts_and_seeds_protein_portion_oz + legume_protein_portion_oz",

"unsaturated_to_saturated_fat_portion_g":

"monounsaturated_fat_portion_g + polyunsaturated_fat_portion_g"

"fatty_acids":

"unsaturated_to_saturated_fat_portion_g / saturated_fat_portion_g"

}

MACRO–CALORIE CONSISTENCY CHECK (required)

After computing totals:

- Compute estimated_calories_from_macros = 4*protein_g + 4*carbohydrates_g + 9*fat_g using TOTALS.

- Enforce consistency: totals.calories_kcal must be within ±10% of estimated_calories_from_macros.

If outside this range, adjust totals.calories_kcal to the nearest value within the ±10% range while leaving macros unchanged.

- Also apply the same consistency check at the INGREDIENT level when possible:

ingredient.calories_kcal should be within ±15% of 4*protein + 4*carbs + 9*fat for that ingredient.

If outside this range, adjust ingredient.calories_kcal to the nearest value within ±15% range while leaving macros unchanged.

UNSATURATED FAT DERIVATION (required)

- If monounsaturated/polyunsaturated are not directly inferable, estimate them using typical composition assumptions.

- Always satisfy: unsaturated_fat_portion_g = monounsaturated_fat_portion_g + polyunsaturated_fat_portion_g.

RETURN ONLY VALID JSON MATCHING THIS SCHEMA EXACTLY

{

"packaged_food_status": "fully_packaged" | "partially_packaged" | "unpackaged",

"ingredients": [

{

"name": "string",

"weight_g": number,

"nutrients": {

"calories_kcal": number,

"fat_g": number,

"carbohydrates_g": number,

"protein_g": number,

"fiber_g": number,

"added_sugars_g": number,

"sodium_mg": number

"monounsaturated_fat_g": number,

"polyunsaturated_fat_g": number,

"unsaturated_fat_g": number,

"saturated_fat_g": number,

}

}

],

"totals": {

"calories_kcal": number,

"fat_g": number,

"carbohydrates_g": number,

"protein_g": number,

"fiber_g": number,

"added_sugars_g": number,

"sodium_mg": number

"monounsaturated_fat_g": number,

"polyunsaturated_fat_g": number,

"unsaturated_fat_g": number,

"saturated_fat_g": number,

},

"hei_components": {

"total_fruit_portion_cup": number,

"whole_fruit_portion_cup": number,

"citrus_melons_berries_whole_fruit_portion_cup": number,

"other_whole_fruits_portion_cup": number,

"fruit_juice_portion_cup": number,

"total_vegetables_portion_cup": number,

"vegetable_legumes_portion_cup": number,

"total_vegetables_and_legumes_portion_cup": number,

"dark_green_vegetables_portion_cup": number,

"greens_and_beans_portion_cup": number,

"whole_grains_portion_oz": number,

"refined_grains_portion_oz": number,

"dairy_portion_cup": number,

"meat_poultry_seafood_protein_portion_oz": number,

"egg_protein_portion_oz": number,

"nuts_and_seeds_protein_portion_oz": number,

"soy_protein_portion_oz": number,

"legume_protein_portion_oz": number,

"high_fat_seafood_protein_portion_oz": number,

"low_fat_seafood_protein_portion_oz": number,

"total_protein_foods_portion_oz": number,

"seafood_and_plant_proteins_portion_oz": number,

}

}

**Supplementary figures and tables**

**Table S1: Multimodal Large Language Models evaluated in phase 1**

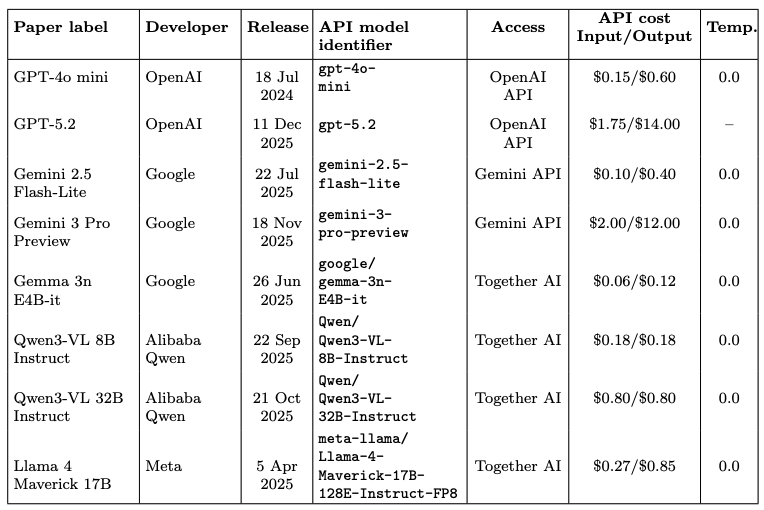

API costs are reported in USD per million input/output tokens. A temperature of 0.0 was used when supported; no temperature parameter was sent for GPT-5.2.

**Table S2: SNAPMe subset comparison of the eight mLLMs with No-CoT and CoT prompting.**

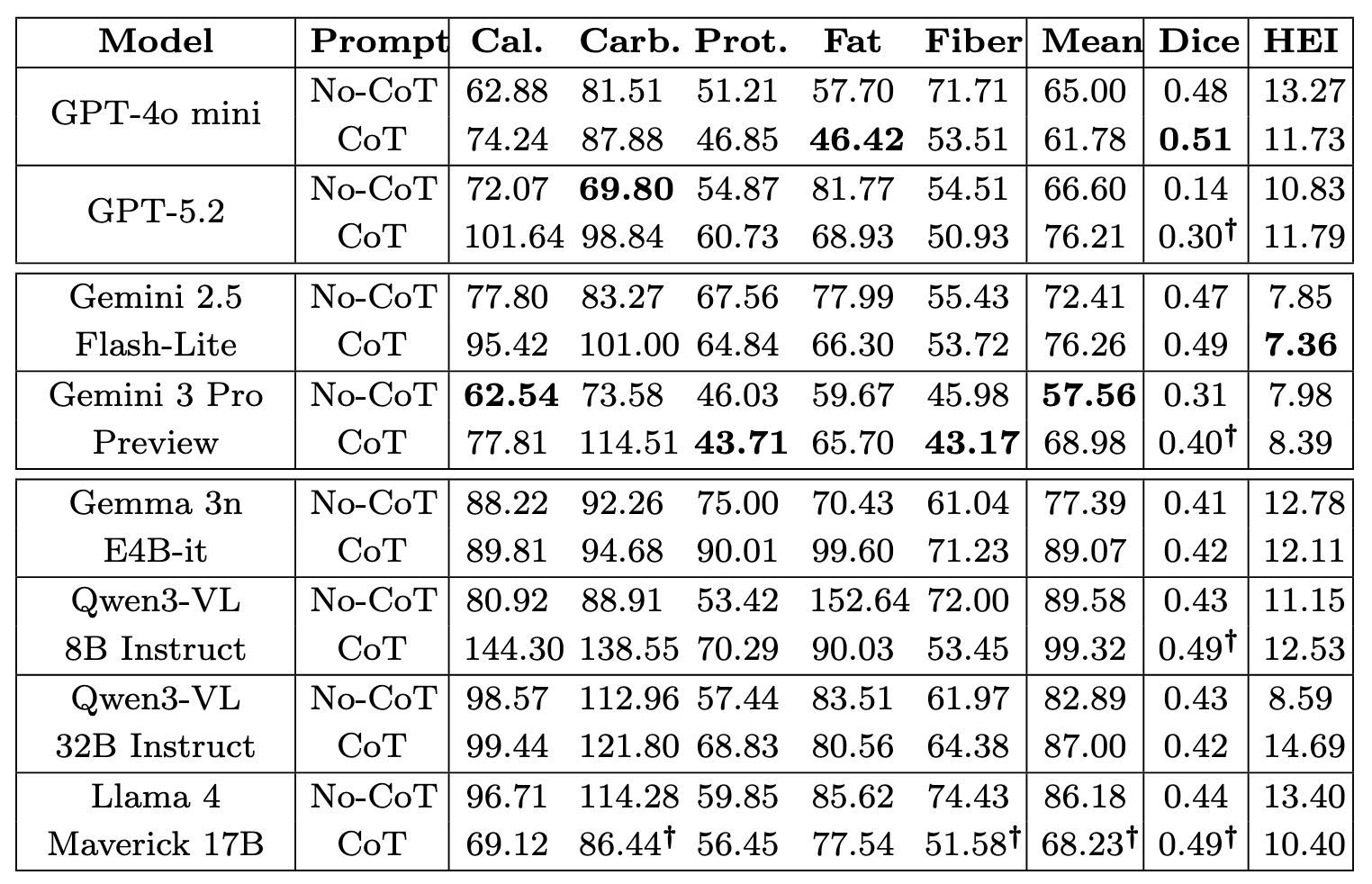

Cal., Carb., Prot., Fat, Fiber, and Mean are MAPE values in percent. Mean is the unweighted mean MAPE across calories, carbohydrates, protein, total fat, and fiber. Dice is the mean Dice score for food item prediction. HEI is RMSE in HEI total-score points. Bold indicates the best value in each column; lower is better for MAPE and HEI RMSE, higher is better for Dice. **†** indicates that CoT was significantly better than No-CoT for that metric within the same model (one-sided paired Wilcoxon signed-rank test for the directional hypothesis that CoT is better than No-CoT, p<0.05).

**Table S3:** **Detailed performance metrics of the three finalist models on the full SNAPMe dataset (1680 photographs; 272 participant-days for HEI), with participant-level bootstrap 95% confidence intervals.**

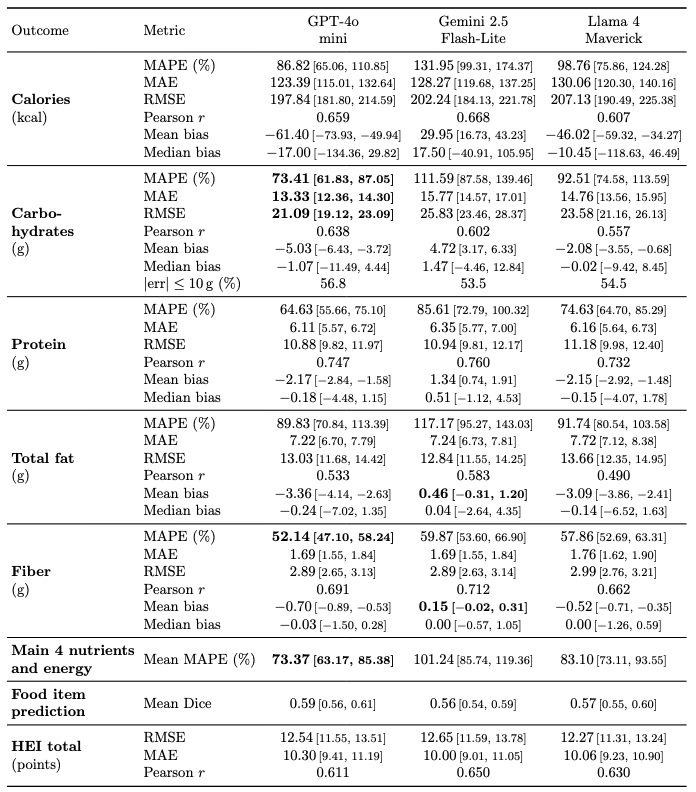

**Brackets after estimates give percentile 95% CIs from 2,000 participant-level bootstrap resamples (seed 42) for nutrient MAPE, MAE, RMSE and mean bias, HEI RMSE and MAE, and mean Dice. All photographs or participant-days from each sampled participant were retained. Mean MAPE is the unweighted mean of the MAPE values for energy, carbohydrates, protein, total fat and fiber, calculated within each bootstrap replicate. For median bias, brackets give the IQR (descriptive, not bootstrapped). Positive bias indicates overestimation. All Pearson correlations were significant at p<0.001. Bold indicates that the best point estimate (closest to zero for bias) was significantly better than both other models, based on participant-level bootstrap CIs of paired differences, Bonferroni-adjusted for the two comparisons per row (97.5% CIs)**

**Table S4. Prediction errors of GPT-4o mini by meal type on the full SNAPMe dataset.**

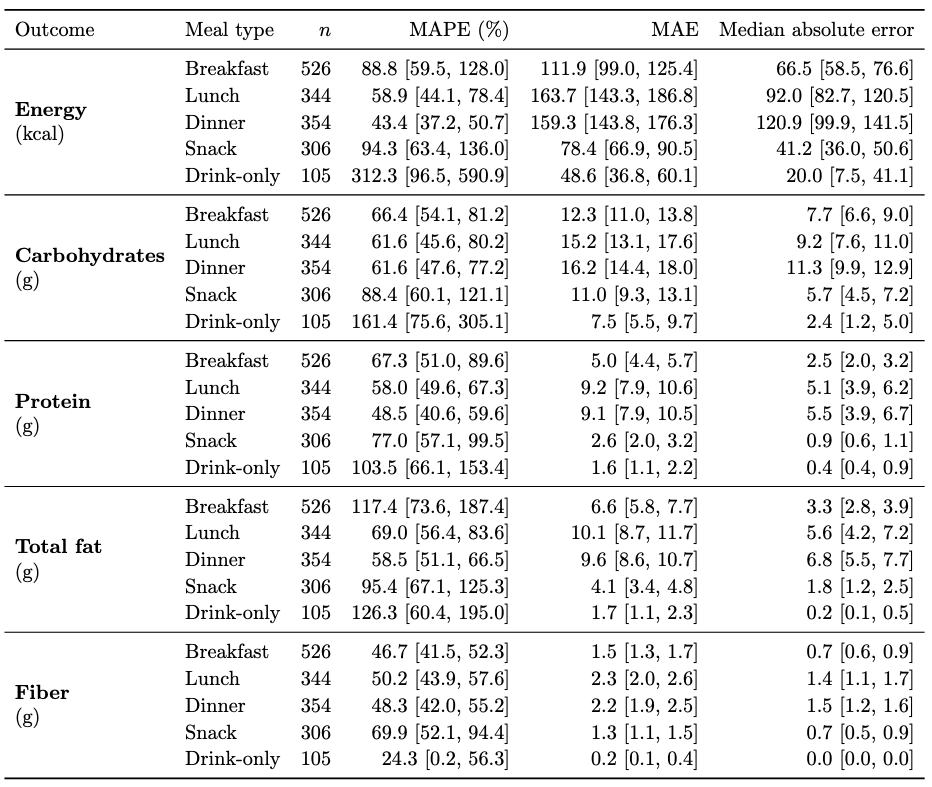

**MAPE, MAE and median absolute error (AE) were calculated from photo-level errors; MAE and median AE are in kcal for calories and in g for nutrients. Brackets give percentile 95% CIs from 2,000 participant-level bootstrap resamples, retaining all eligible photos of each sampled participant. MAPE follows the near-zero rule described in SM1. Photographs labeled as supper or brunch were excluded.**

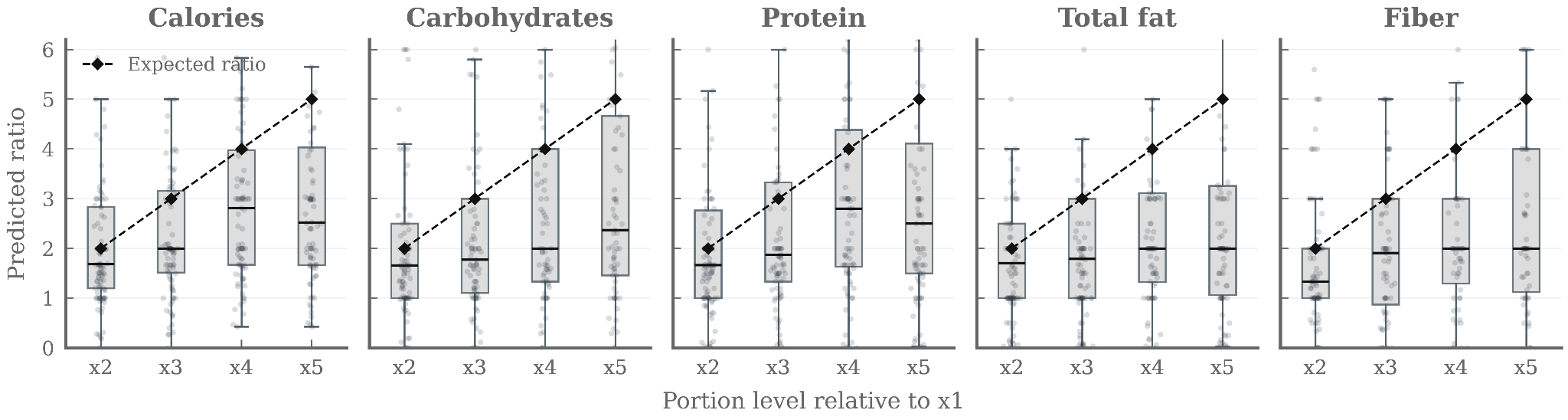

**Figure S1. Portion-size sensitivity of nutrient and energy predictions. Predicted values for calories, carbohydrates, protein, total fat, and fiber are expressed as ratios relative to the smallest reference portion (×1) across 100 food items from the ASA24 portion library.**

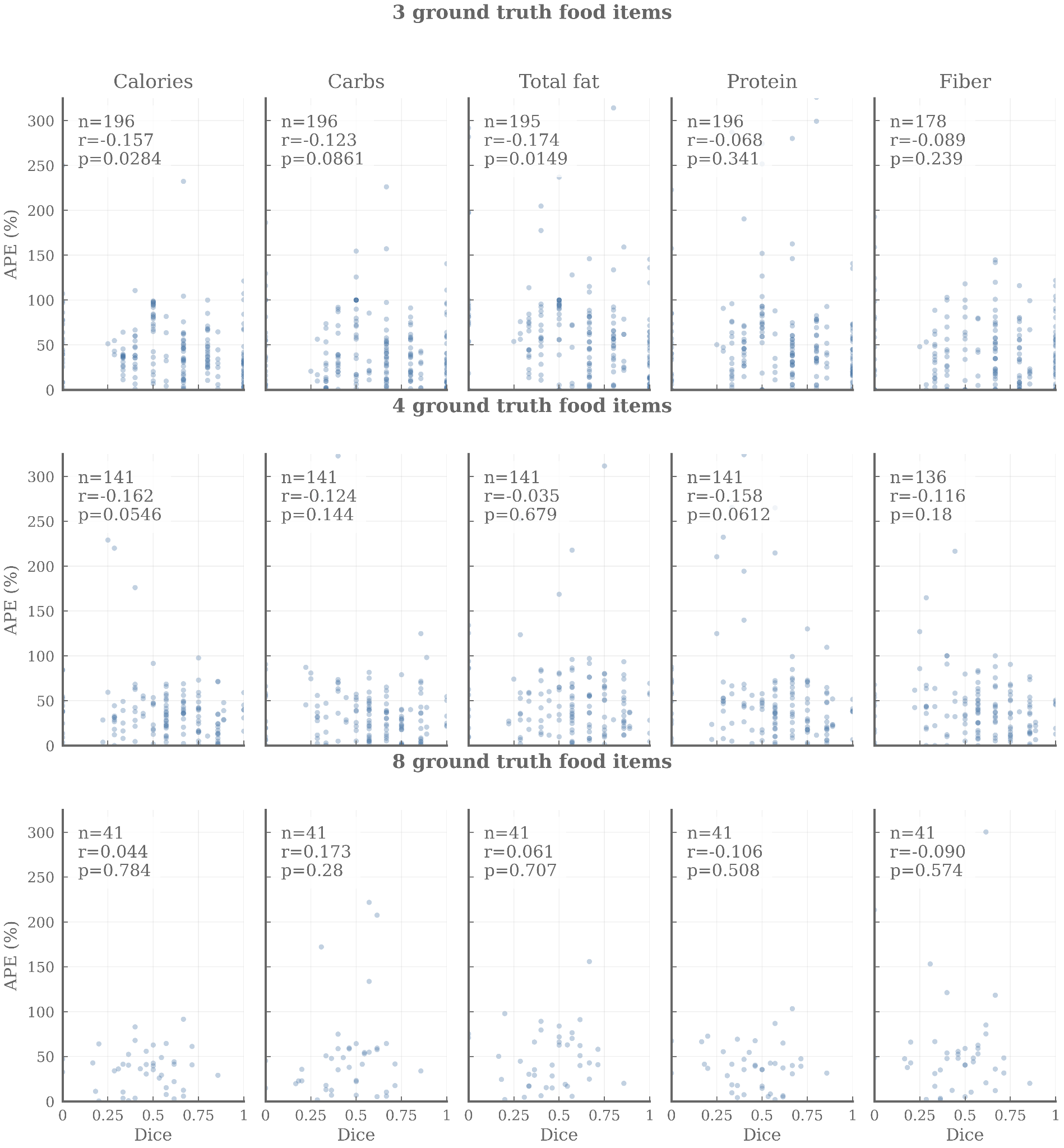

**Figure S2: Association between food-item recognition accuracy and nutrient prediction errors by ground-truth food item count.**
